# *Healthy Heart Actions Right Time* (HHART): Co-design priorities to connect Aboriginal and Torres Strait Islander community and clinic activities for healthy hearts

**DOI:** 10.64898/2026.06.05.26354870

**Authors:** Rosemary Wyber, Jonathon Zagler, Chelsea Liu, Uday N. Yadav, Zoe O’Dwyer, Kirsty Hart, Kane Chapman, Lisa McGrady, Alicia Kohn, Narelle Winterfield, Dylan Williams, Nathan Watson, Kim Morey, Odette Pearson

**Author notes:** Corresponding author: Dr Rosemary Wyber, OR Prof Odette Pearson.

## Abstract

**Aim:** Healthy Heart Actions Right Time (HHART) is a multi-phased research project that seeks to identify, implement and evaluate strategies to connect community and clinical activities to reduce the burden of heart disease for Aboriginal and Torres Strait Islander people. The aim in Phase One was to identify priority activities for two participating services.

**Background:** The ongoing effects of colonisation drive a disproportionate burden of heart disease for Aboriginal and Torres Strait Islander people. Clinical and community groups both have established strengths in reducing the risk of heart disease, but these are not always well connected.

**Methods:** Using a case study methodology in two locations we partnered in a 12-month co-design process to identify priority activities to connect clinical and community activities.

**Findings:** Three priorities emerged from the Phase One co-design process: (i) community-led gardening as a strategy to promote heart health through connection and healthy lifestyles; (ii) community days to increase engagement in heart checks and strengthen community-clinic relationship; and (iii) clinic-led development of culturally relevant education resources to promote clinician confidence and community heart health knowledge.

## Introduction

Aboriginal and Torres Strait Islander peoples, the First Peoples and sovereign custodians of Australia, are leaders in collective approaches, activities and advocacy that drive action for health and wellbeing. Community-based approaches have been critical in the success of the Aboriginal community-controlled primary health services movement, emergency responses and aged care [1–4]. Community leadership also offers a powerful approach to address the disproportionate burden of chronic disease for Aboriginal and Torres Strait Islander people [5]. Heart disease in the Aboriginal and Torres Strait Islander population occurs at twice the rate of the non-Indigenous population and has a particular presentation of younger onset and disproportionately poor long-term outcomes [6]. This burden is driven by the effects of colonisation, racism and socioeconomic marginalisation [7]. Similar patterns of cardiovascular inequity have been documented among Indigenous populations worldwide, where colonisation, structural inequities and barriers to culturally safe care continue to shape cardiovascular outcomes [8, 9]. It follows that community-led approaches to improving heart health, which include attention to these drivers, are important for impact on outcomes.

Within Australia, the concept of comprehensive integrated approaches to reducing heart attack and stroke has been developing for decades. It is enshrined in jurisdictional and national policy documents, including the South Australian Heart and Stroke Plan 2022-2027 [10], the National Aboriginal and Torres Strait Islander Health Plan 2021-2031 [11] and the National Strategic Action Plan for Heart Disease and Stroke [12]. In the context of Aboriginal and Torres Strait Islander health, comprehensive integrated approaches extend beyond biomedical models of prevention and treatment to include culturally grounded, community-led and holistic approaches to health and wellbeing [13–15]. Despite increasing policy recognition of integrated and holistic models of care, there has been relatively little evidence-informed work on how to operationalise community and clinic level activities to improve heart hearth. Community capabilities may be overlooked in the design and delivery of heart health programs, which can result in a narrow clinical and biomedical focus. This challenge is not unique to Australia, with international literature similarly identifying gaps between Indigenous health policy aspirations and the implementation of culturally informed cardiovascular prevention approaches in practice [16].

The Healthy Heart Actions Right Time (HHART) project was established to explore how holistic and culturally meaningful approaches to heart health can be integrated into clinical activities and community business. This paper outlines the processes and outcomes of Phase One – a co-design journey with partner primary care services in two locations in Australia. Accordingly, this manuscript reflects the methods through which activities were collectively developed, including partnership development, planning, co-design workshops, and iterative refinement.

## The Co-Design Approach

### Study Design and Approach

The need for HHART was initially identified during collaborative work between research teams from the Wardliparingga Aboriginal Health Equity Theme at the South Australian Health and Medical Research Institute and the Yardhura Walani Centre for Aboriginal and Torres Strait Islander Wellbeing Research at the Australian National University. Through this collaboration, it became evident that while recommendations for heart health frequently emphasised ‘community engagement’, there were few well described examples of this in practice. HHART was subsequently funded by the Medical Research Future Fund Targeted Call for Research on Cardiovascular Health (GNT: 2032711). The grant was co-written with three partner organisations, two of whom remain in the project as described in this study and one who withdrew, once the grant was awarded, due to competing workforce demands and shortages.

The broader HHART research project is a mixed-methods program of research designed to strengthen connections between community and clinic approaches to heart health. The project comprises two phases, with Phase One focused on the co-design of locally relevant activities and implementation approaches, and Phase Two focusing on implementation and ongoing evaluation of these activities. Phase One followed a multiple case study approach [17, 18] underpinned by co-design principles [19, 20]. We drew upon the principles outlined by Anderson et al., (2022) [Table 1] to enable in-depth exploration of local contexts and priorities across two participating Aboriginal community-controlled health organisations, Indigenous Wellbeing Centre (IWC) and Pangula Mannamurna Aboriginal Corporation (PMAC). IWC is located in Bundaberg, Queensland. IWC services over 16,500 clients across the traditional lands of the Taribelang Bunda, Gooreng Gooreng, Gurang and Bailai Peoples and delivers a broad range of clinical, social and cultural services [21]. PMAC operates on Boandik Country (Mount Gambier, South Australia). PMAC services between 800-1000 clients across the Limestone Coast, including primary care, outreach services, and holistic support programs [22].

**Table 1.**
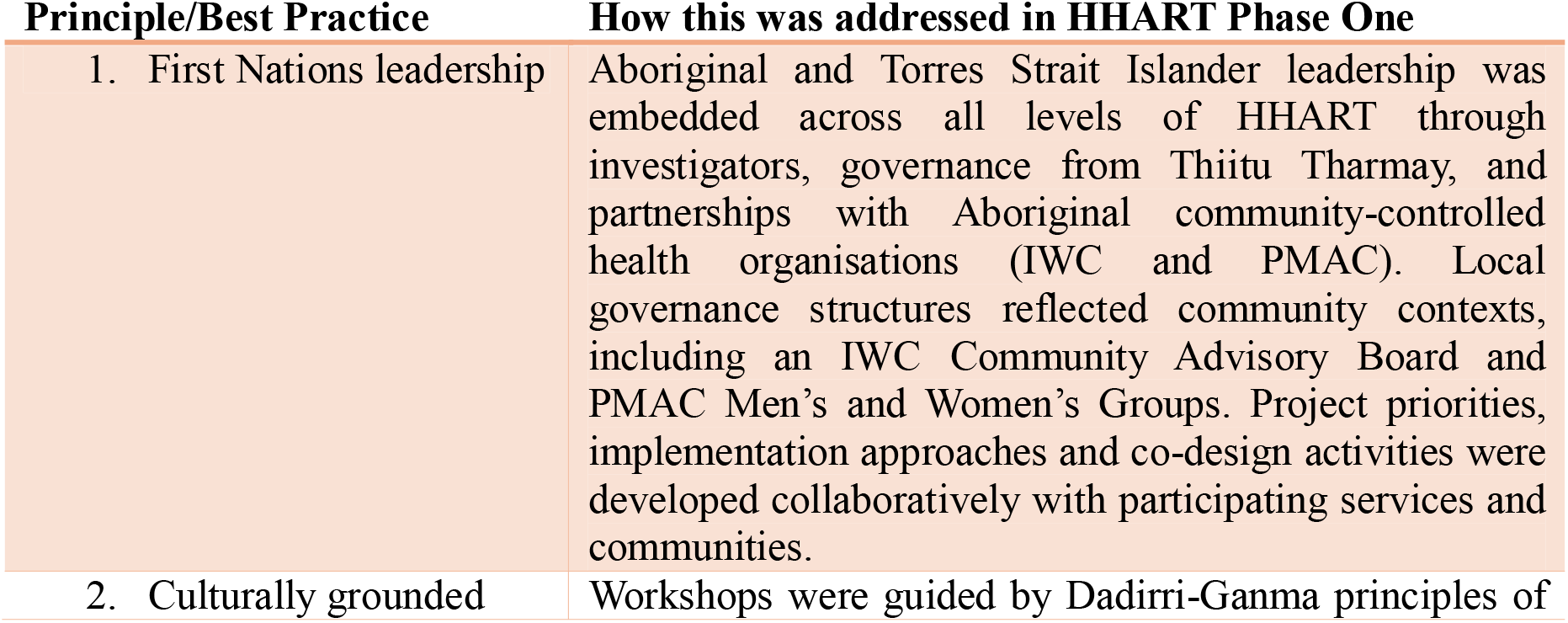
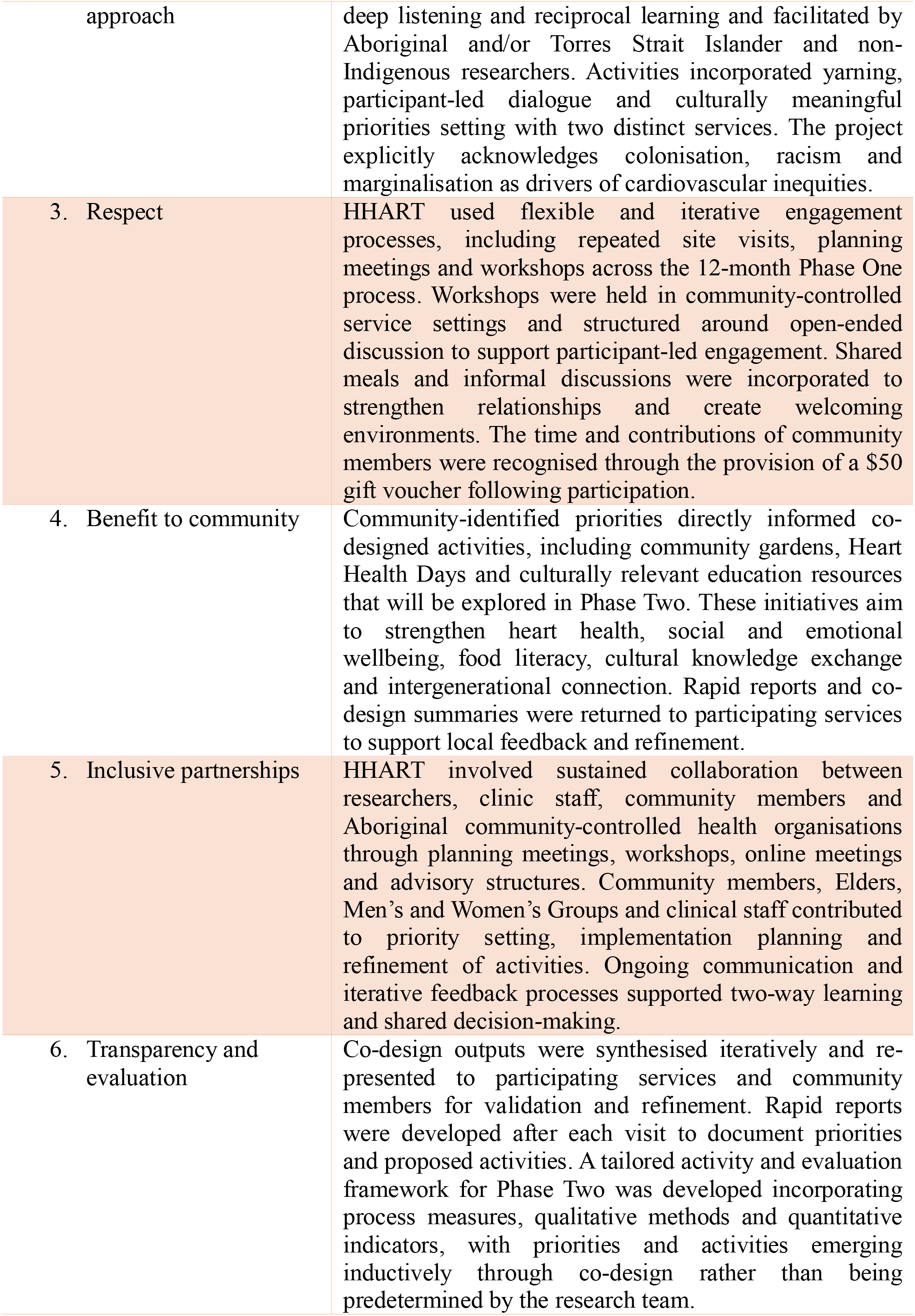
Co-design principles [19].

Co-design was undertaken as an iterative and relational process across all stages of Phase One [Figure 1]. Insights generated through planning meetings, workshops and informal engagements were continuously reflected on, refined and reintroduced in subsequent activities. This enabled the progressive development of program components that were responsive to local contexts, priorities and Aboriginal ways of knowing, being and doing.

**Figure 1.**
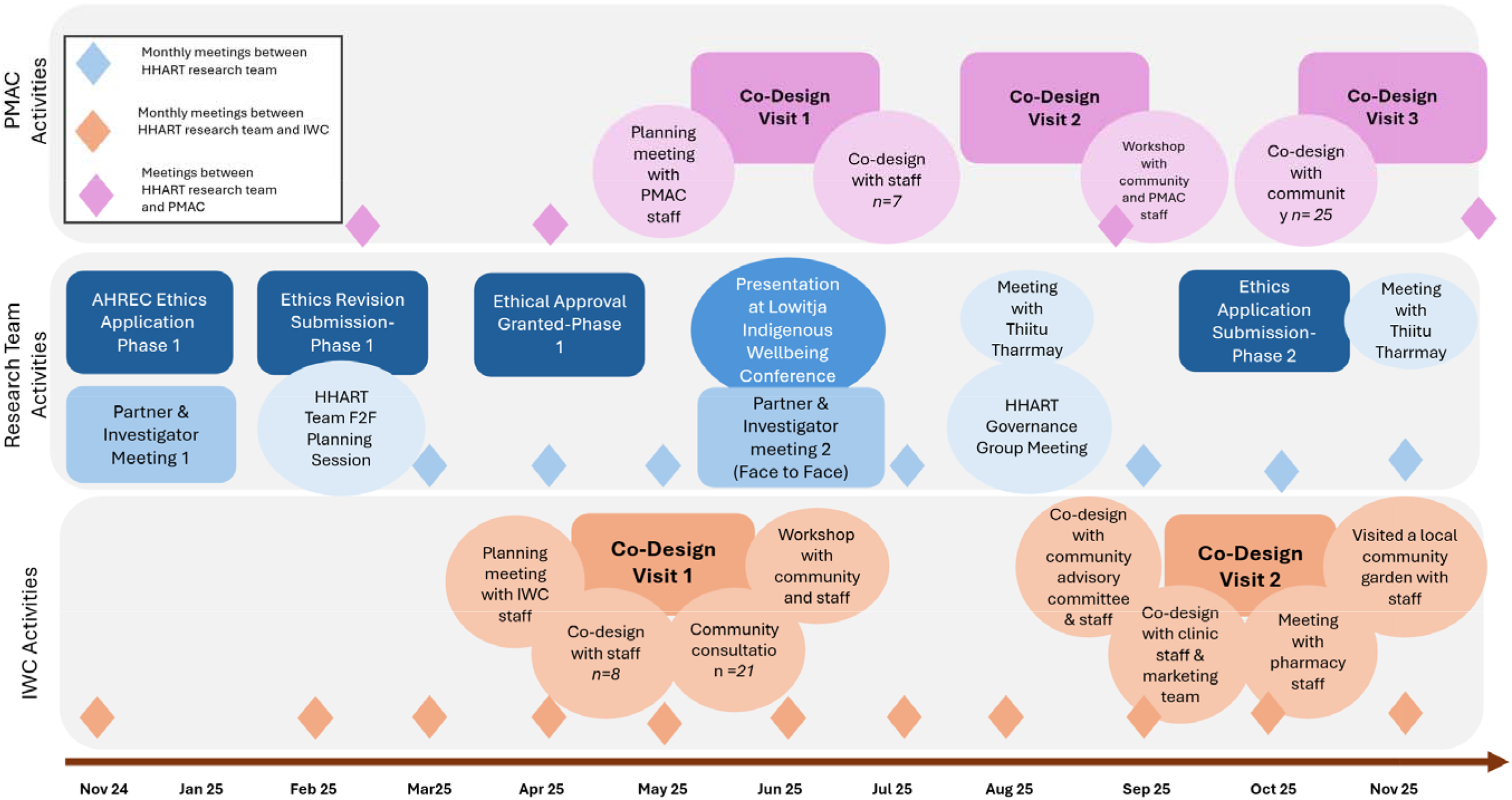
Co-design overview.

### Ethics and Governance

Ethical approval to conduct Phase One was granted by the South Australian Aboriginal Health Research Ethics Committee (AHREC) (#04-24-1160). Throughout the research process, the nine ethical principles of conducting research with Aboriginal and Torres Strait Islander communities in South Australia, as outlined in the South Australian Aboriginal Health Research Accord [23], were followed.

HHART is underpinned by a multidimensional governance and operational framework that enshrines Aboriginal and Torres Strait Islander leadership and sovereignty at all levels, outlined in Figure 2. Thiitu Tharrmay (Ngiyampaa language, translating to “to share/exchange knowledge”) is the Aboriginal and Torres Strait Islander Reference Group at Yardhura Walani, Australian National University [24]. The HHART research team engaged with Thiitu Tharrmay at inception and during project development with presentations and feedback in August 2023, August 2024 and November 2025. A governance planning meeting was held with participating services in August 2025, culminating in a plan for local input via Community Advisory Groups and connection across the project as required through the ‘HHART Beat’ network. At IWC, governance is provided by clinic staff (KH, KC, LM, AK) and a community advisory board established for the purposes of the HHART project. At PMAC, governance is led by clinic staff (NW, DW, NW) in partnership with existing Men’s and Women’s Groups. Additionally, the HHART Investigator group, comprising a majority Aboriginal and/or Torres Strait Islander peoples, bring leadership and direction grounded in Indigenous philosophies. The Investigator group has met twice during Phase One, supported by monthly working team meetings of project co-leads (OP, RW) and research staff (JZ, CL, ZO).

**Figure 2.**
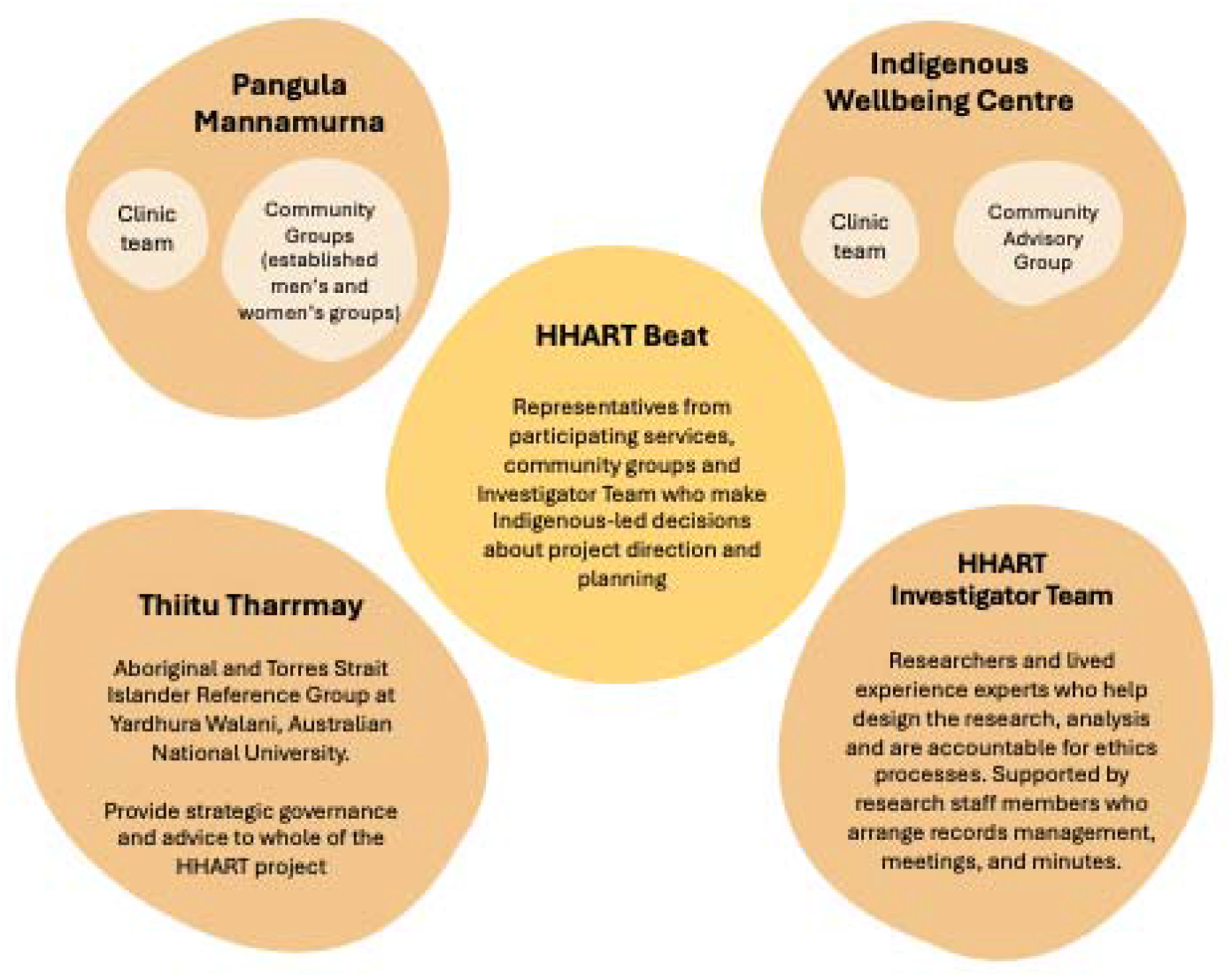
HHART Beat governance structure.

### Iterative Development and Refinement

Thorough preliminary planning meetings and email consultations between the HHART research team and participating services occurred, with co-design activities scoped and refined in partnership with each site. These meetings were a critical component of the co-design process, providing a space for relationship building, shared decision-making, and alignment of expectations prior to formal workshops. While not treated as formal data collection, they enabled the research team and services to identify locally relevant priorities, clarify roles, and ensure that subsequent co-design activities were responsive to community and service contexts. These engagements supported alignment between project aims, local priorities, and service contexts, ensuring that co-design activities were grounded in community-identified needs. Across both sites, planning activities were relational and iterative, with ongoing dialogue between community members, clinic staff, and the research team shaping the direction and focus of the co-design process.

At IWC, the first site visit in May 2025 commenced with a planning meeting attended by IWC staff and Board members alongside the research team (RW, UYN, OP). This meeting focused on confirming project aims, identifying local priorities, and considering implementation approaches within the service context. A second three-day visit to IWC in October 2025 focused on refining program implementation strategies (RW, CL, JZ). This included consultation with the IWC Community Advisory Group, who provided feedback to inform the next phase of the project, as well as meetings with IWC staff, communications personnel, and pharmacy staff. These engagements supported the development of program implementation, communication strategies, and opportunities to strengthen medication-related health information. A workshop with IWC staff further explored the development of a health assessment workflow and program evaluation plan.

Similarly, at PMAC, planning processes occurred iteratively alongside and between co-design workshops. Two in-person planning workshops were held in April 2025 (OP, KM) and July 2025 (OP, JZ), complemented by regular online meetings with clinic staff to support continuity and coordination. The initial April workshop focused on establishing shared understandings of the project and exploring early priorities, while a second workshop in July 2025 engaged community members in further identifying and refining priorities and potential activities. Between these time points, ongoing discussions with clinic staff enabled the research team to remain responsive to local contexts and emerging needs. In addition, an informal planning meeting between PMAC staff and the research team following the initial workshop supported the refinement of subsequent co-design activities and ensured that project directions were aligned with community-identified priorities. A final meeting was held in April 2026 to finalise co-design outcomes.

### Co-design Workshops

Co-design workshops formed the primary mechanism for collaboratively developing program activities. Following initial planning, a series of in-person workshops were conducted with each participating service. This included *n=2* with IWC and *n=2* with PMAC.

People were invited to participate in structured focus groups to provide systematic input into designing the project activities. Participants were recruited using a community-led approach that combined purposive sampling and open community invitation. Purposive sampling was used to engage clinic staff and Aboriginal and Torres Strait Islander peoples receiving services through the participating health services. All participants were 18 years or older, there were no exclusion criteria for people who wished to be involved. Staff participants were nominated and approved by each health service to contribute to project development, and established community groups, such as Elders’, Men’s, and Women’s groups, were invited to participate in co-design activities. To support recruitment, study flyers developed by the research team were circulated through the participating health services to invite community members and relevant groups to take part.

Workshops were informed by a collaborative, community-based approach [25] and guided by principles of Dadirri-Ganma (deep listening and reciprocal learning), with each session commencing with time for introductions and yarning to support relationship building and shared understanding [26] [Figure 3]. Workshops were held in private spaces provided by each service and guided by open-ended discussion frameworks to enable flexible, participant-led dialogue.

**Figure 3.**
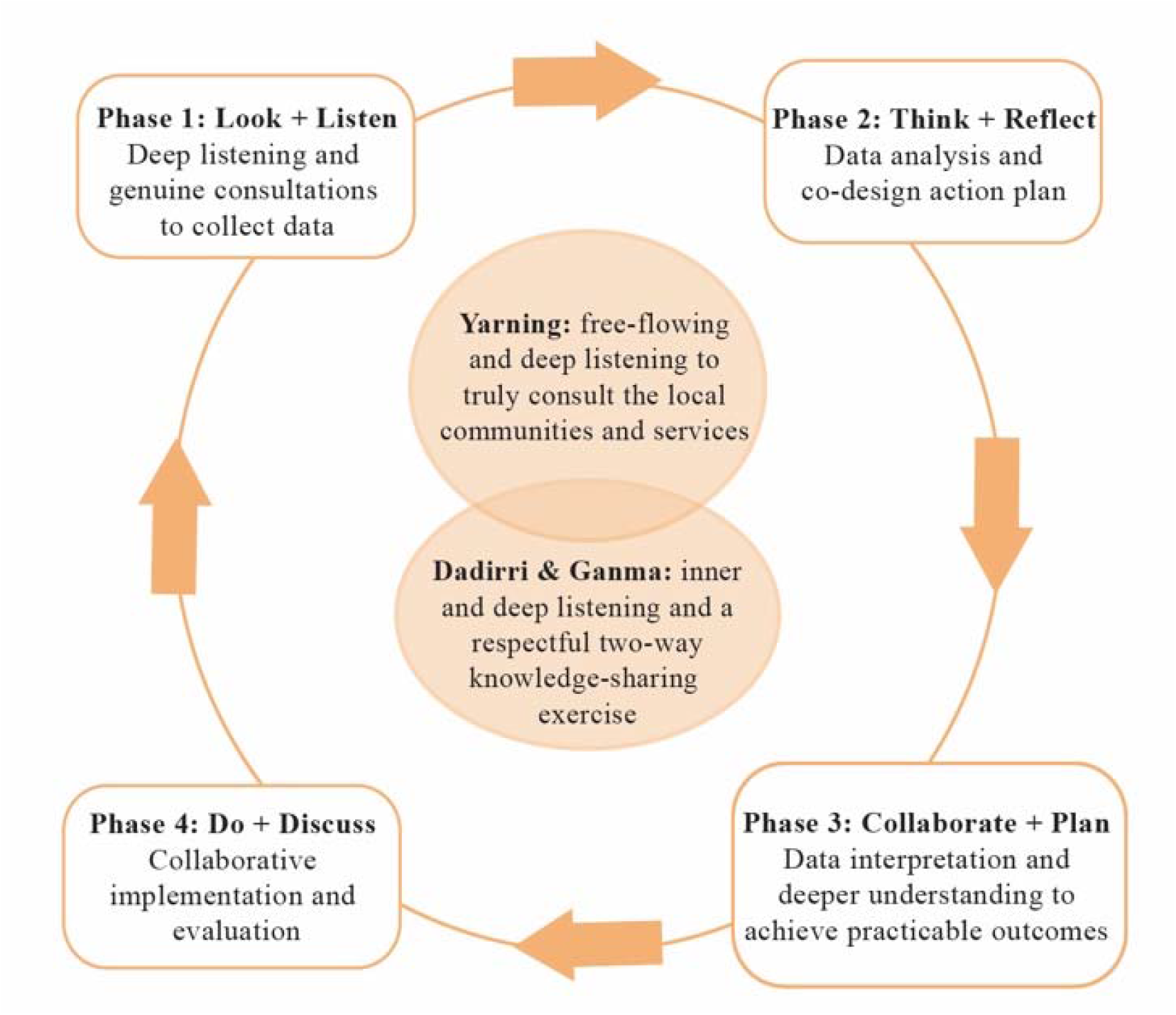
PAR-Dadirri-Gamma approach, adapted from Sammil et al. 2021.

During the first visit to IWC, a series of co-design workshops were held with both community members and staff. A community workshop involving 23 participants explored local perspectives on heart health priorities and potential program activities facilitated by Aboriginal and/or Torres Strait Islander and non-Indigenous members of the research team. A staff-only workshop with seven clinical team members focused on service delivery considerations and feasibility. Insights from these sessions were reviewed and synthesised by the research team between workshop days and subsequently discussed in a triangulation workshop involving both staff and community members to identify shared priorities and areas of alignment. Workshops were jointly facilitated by Aboriginal and/or Torres Strait Islander (OP) and non-Indigenous (RW, CL) members of the research team.

At PMAC, a series of co-design workshops were conducted between June and November 2025. The first workshop, held in June 2025, focused on eliciting perspectives from clinic staff and was attended by seven participants, followed by a further community-focused workshop in November 2025 with 25 participants. Workshops were jointly facilitated by Aboriginal and/or Torres Strait Islander (OP, KM) and non-Indigenous (JZ) members of the research team. Reflecting the participatory nature of the approach, butchers’ paper was used to capture participant ideas, reflections and priorities throughout sessions, with these materials collected as part of the co-design outputs. At PMAC, workshops were complemented by visits to significant places within the service environment, further grounding discussions within local cultural and physical contexts.

Across both sites, workshops concluded with shared meals, providing opportunities for informal discussion and strengthening relationships between participants and the research team. With participant permission, IWC workshops were audio recorded. PMAC workshops were not audio-recorded, with facilitator notes and participant-generated materials used to document discussions.

### Interpretation of Co-Design Outputs

Co-design outputs were synthesised using a co-design-informed multiple case study approach, with each service treated as a distinct case reflecting its local community and organisational context [17, 18]. Data from each site were interpreted separately to preserve local specificity. For each site, all co-design data (e.g., workshop discussions, facilitator notes) were compiled and reviewed to develop an overall understanding of context and priorities.

Consistent with co-design principles [27], analysis was iterative and inductive, focusing on identifying participant-defined priorities, program components and implementation and evaluation considerations. Interpretations were refined across successive workshops as co-design and planning progressed. Analysis was led by RW, UNY and CL for IWC, and OP and JZ for PMAC, with all team members contributing to reflection and refinement of findings, including synthesis to community members for validation.

Analytic outputs were used to directly inform program design and implementation. Rapid reports (case summaries) were developed to maintain a clear link between participant input and the resulting co-designed activities and decisions, supporting transparency and rigour. After each visit, a rapid report was provided to each service to share findings and invite further feedback [Supplementary File 1].

### Co-Design Outcomes

The findings presented in this section are drawn from co-design workshops involving formally recruited and consenting participants. While the broader co-design process included planning meetings, informal engagements and ongoing collaboration with services, these interactions are not reported as data within this analysis. Rather, they informed the iterative development of activities and provided contextual grounding for the findings presented here. Illustrative quotes for IWC are presented in Table 2. As PMAC co-design workshops were not audio recorded, no quotes are provided.

**Table 2.**
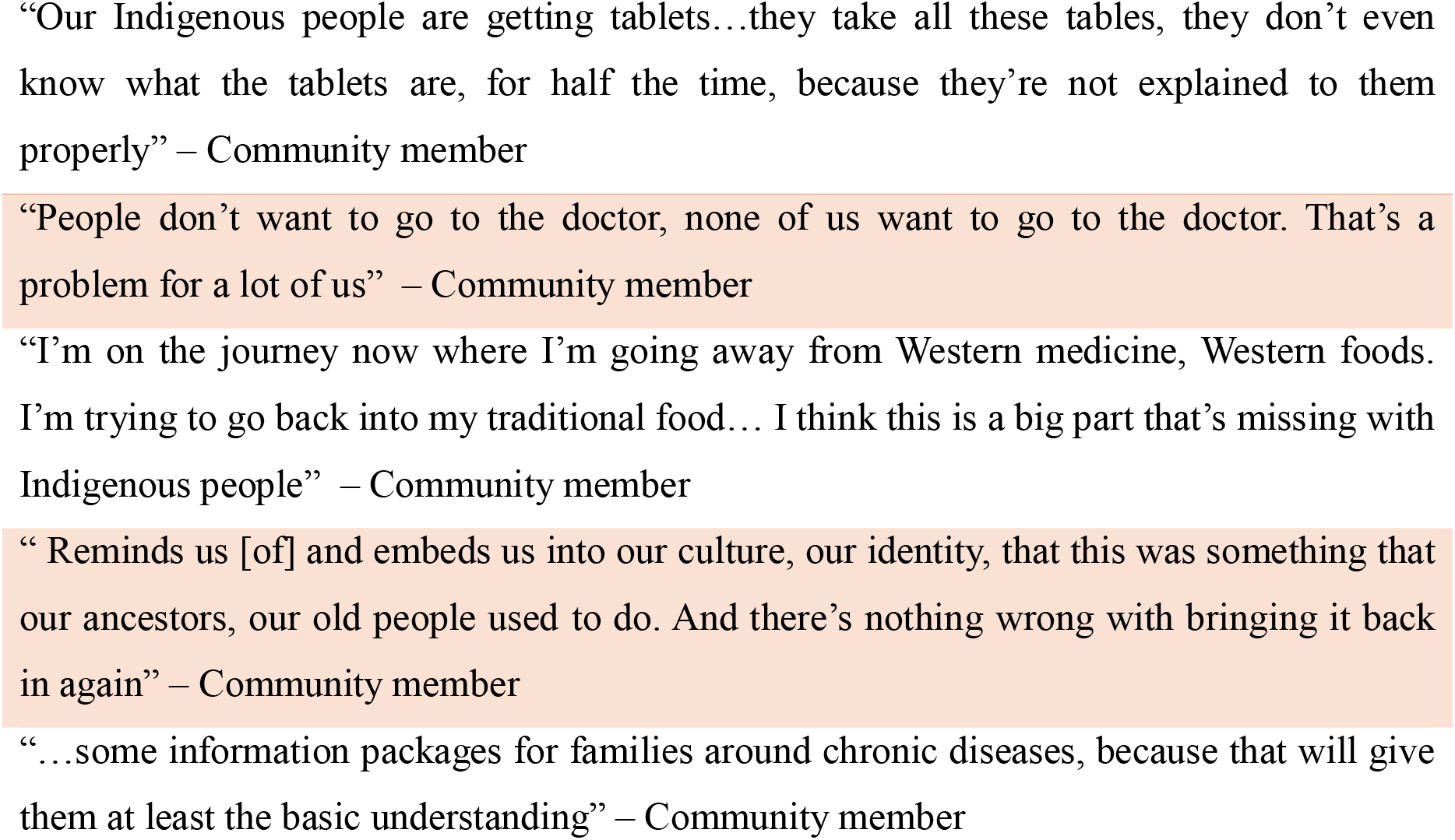
IWC illustrative quotes.

### Site One – IWC

Feedback during the community workshop revealed strong interest in preventing heart disease, in addition to treatment for people with established disease. There was a sense that heart disease causes too much sickness and suffering and that people wanted to ‘take control’ of the effects of heart disease. Participants expressed a strong interest in use of traditional foods and cultural medicines as a way of achieving this and keeping hearts strong.

Above all there was clear pride and comfort in traditional food and medicine. Some people wanted to be able to share this cultural knowledge with clinical staff and suggested opportunities to share traditional food with staff or talk with practice staff in non-clinical contexts. In contrast to the enthusiasm for traditional medicines participants reflected that they felt they understood biomedical/clinical care not as well, with some expressing confusion or frustration with medication and medical care. Others spoke positively about the good care they had experienced, particularly for serious heart issues. Variability in care experiences was widely acknowledged, often dependent on a particular doctor or clinician. In general, heart health care in primary care was more positively regarded than heart health interactions with hospital or specialist care clinicians.

There was a sense that participants wanted more information about heart health to inform their own understanding and to better support family members in preventing heart disease or living with heart disease. This also included a desire for yarning about heart health, particularly with Aboriginal Health Workers/Practitioners, and accessible resources such as information on TV screens in waiting areas and take-home information on heart health. People wanted to feel prepared when they were meeting with a specialist. There was a strong focus on collective approaches with participants identifying families, group activities and community events as potential approaches to support heart health. Some people identified that events, activities and outreach provide an opportunity to engage more young people in heart health conversations.

Content in the clinical staff focus group spontaneously included some similar themes. Staff wanted more opportunities to connect and yarn in non-clinical settings with community members and current clients and identified several previous activities which had supported this, including outreach days, events in parks and walking groups. There was talk about ‘mums and bubs’ groups, women’s groups and sports teams as outreach opportunities. When discussing different initiatives and opportunities clinical participants often said that they ‘had done that in the past’ or ‘always wanted to do that’. There was a sense that young people like hearing from Elders and that there were opportunities to better engage young people in health care.

Clinical staff also reflected on their health promotion experiences, identifying that resources people could ‘touch and feel’ were well received. For example, a previous interactive model of cholesterol plaques narrowing cardiovascular arteries was fondly recalled. Staff wanted people to be able to interact with education resources independently and in their own time, such as with interactive touch screens or 3D models in waiting areas.

### Site Two – PMAC

Across community and clinic staff workshops, participants articulated a shared, holistic understanding of heart health that extended beyond clinical care to encompass physical activity, cultural connection, social relationships and access to services. Heart health was consistently framed as something embedded in everyday community life and shaped by cultural, social and structural conditions.

Gardening emerged as a central strategy for promoting heart health. Participants described gardening as an accessible and culturally meaningful form of physical activity that supports social connection, food security and social and emotional wellbeing (SEWB). Community members emphasised gardening as a space for learning, sharing food and sustaining engagement with the health service, while the clinic staff highlighted the long history of intergenerational gardening within the local community. However, participants noted that previous garden initiatives had been difficult to sustain without ongoing resourcing.

Health education was viewed as most effective when delivered through practical, skills-based activities. Community members proposed regular heart health activities and events that combined hands-on learning with guest speakers (e.g., physiotherapists, dieticians) to build health literacy, confidence and relationships between clinicians and community members. Clinic staff emphasised addressing key heart disease risk factors, including smoking, vaping, nutrition and hidden sugars, and highlighted oral health as a critical but often overlooked contributor to heart disease.

Intergenerational connection and participation were identified as foundational to prevention. Both groups emphasised the importance of involving children, families and Elders in shared activities such as walking groups, gardening, outings on Country and community events. These activities were seen as promoting physical activity, strengthening belonging and supporting positive mental health, while also embedding healthy habits early in life and reinforcing collective responsibility for wellbeing.

Cultural knowledge and connection to Country were central to sustaining wellbeing. Participants emphasised the integration of bush foods, bush medicine and traditional plant knowledge into health activities, viewing cultural knowledge exchange as both preventative and restorative. Opportunities for Elders and young people to learn together were considered essential to maintaining cultural continuity, identity and pride.

Finally, clinic staff highlighted significant structural barriers to heart health care, including limited access to General Practitioners, dental services and specialist care, challenges completing the Aboriginal and Torres Strait Islander Health Assessments (715s), and constraints associated with travel and telehealth access. Here, the importance of community-based preventative approaches that reduce reliance on fragmented clinical services was identified.

## Discussion

The findings from Phase One of HHART highlight the importance of strengthening the connections between community-led activity and clinical care in efforts to prevent cardiovascular disease among Aboriginal and Torres Strait Islander peoples. Participants across both sites consistently framed heart health as something embedded in everyday life, not only shaped by biomedical risk factors but also by the interplay of social relationships, cultural practices, opportunities for physical activity, and access to culturally safe services. These findings reinforce a growing body of literature, both within Australia and internationally, demonstrating that effective chronic disease prevention requires approaches that extend beyond biomedical models of care to encompass the social and cultural contexts in which health is lived [28–30]. Similar findings have been reported across Indigenous health settings internationally, where community-led and relational models of care have been identified as critical to improving engagement, trust and continuity of cardiovascular prevention and care [31].

Importantly, this study demonstrates the potential of community- and clinic-led activities when they operate in synergy. While primary health care services play a critical role in screening, treatment and management of cardiovascular risk [32], participants emphasised that prevention is most effective when it is embedded in community settings where people gather, learn, and support one another. In this sense, community-based initiatives function as important extensions of clinical care, enabling prevention efforts to reach beyond the consultation room and into the everyday spaces where health behaviours and social norms are shaped. These findings align strongly with the long-standing principles of comprehensive primary health care that underpin the Aboriginal community-controlled health sector and have demonstrated the effectiveness of integrating clinical services with Aboriginal and Torres Strait Islander ways of knowing, being and doing [4, 14].

Findings from the co-design workshops have direct implications for the development and implementation of locally responsive heart health initiatives. A shared finding across both Phase One sites was that community and clinic partners independently identified the establishment of a community garden as a locally meaningful priority for supporting heart health and wellbeing. This convergence strengthened the rationale for making community gardening a key initiative for implementation and evaluation in in Phase Two of HHART. Each service’s HHART program will centre on the establishment of a community garden located on site, designed to function as both a practical and *symbolic* hub for heart health promotion. The aspirations articulated during co-design were for the garden to support regular, structured activities that promote physical activity, intergenerational participation, and food literacy. It will provide opportunities for hands-on learning, including growing fruit and vegetables, integrating traditional foods where appropriate, and cooking and nutrition workshops using harvested produce. Importantly, the garden is intended not only as a health intervention but as a relational space that fosters conversation, cultural knowledge exchange, and sustained community engagement. Building on participants’ recommendations, the program will also incorporate recurring *Heart Health Days* featuring guest speakers such as dieticians, exercise physiologists, pharmacists, and oral health professionals. These events will prioritise practical, skills-based education addressing key heart health risk factors. Tailored sessions will be embedded within existing community groups (e.g., youth and Elder’s groups). Additionally, the development of culturally relevant heart health resources will support ongoing education beyond structured events. While these findings are specific to each health service, collectively they highlight the potential for community-based prevention initiatives to operate as scalable and adaptable approaches across diverse Indigenous primary health care settings.

To facilitate this, co-design insights have been developed into a tailored activity and evaluation framework that integrates community-led, clinic-led and partnership-based approaches to heart disease prevention [Figure 4]. Importantly, these activities are not conceived as discrete interventions, but as interconnected components of a broader system designed to strengthen engagement between community members and clinical services. The accompanying evaluation plan reflects this integrated approach, combining process measures (e.g., participation rates, service utilisation, resource engagement) with qualitative methods (e.g., photovoice, focus groups, yarning) and quantitative metrics (nKPI data) to capture both reach and lived experience. This broad evaluation will enable the assessment of not only whether activities are delivered, but how they are experienced and sustained within community contexts, generating practice-based evidence that may inform future Indigenous cardiovascular prevention initiatives both within Australia and internationally.

**Figure 4.**
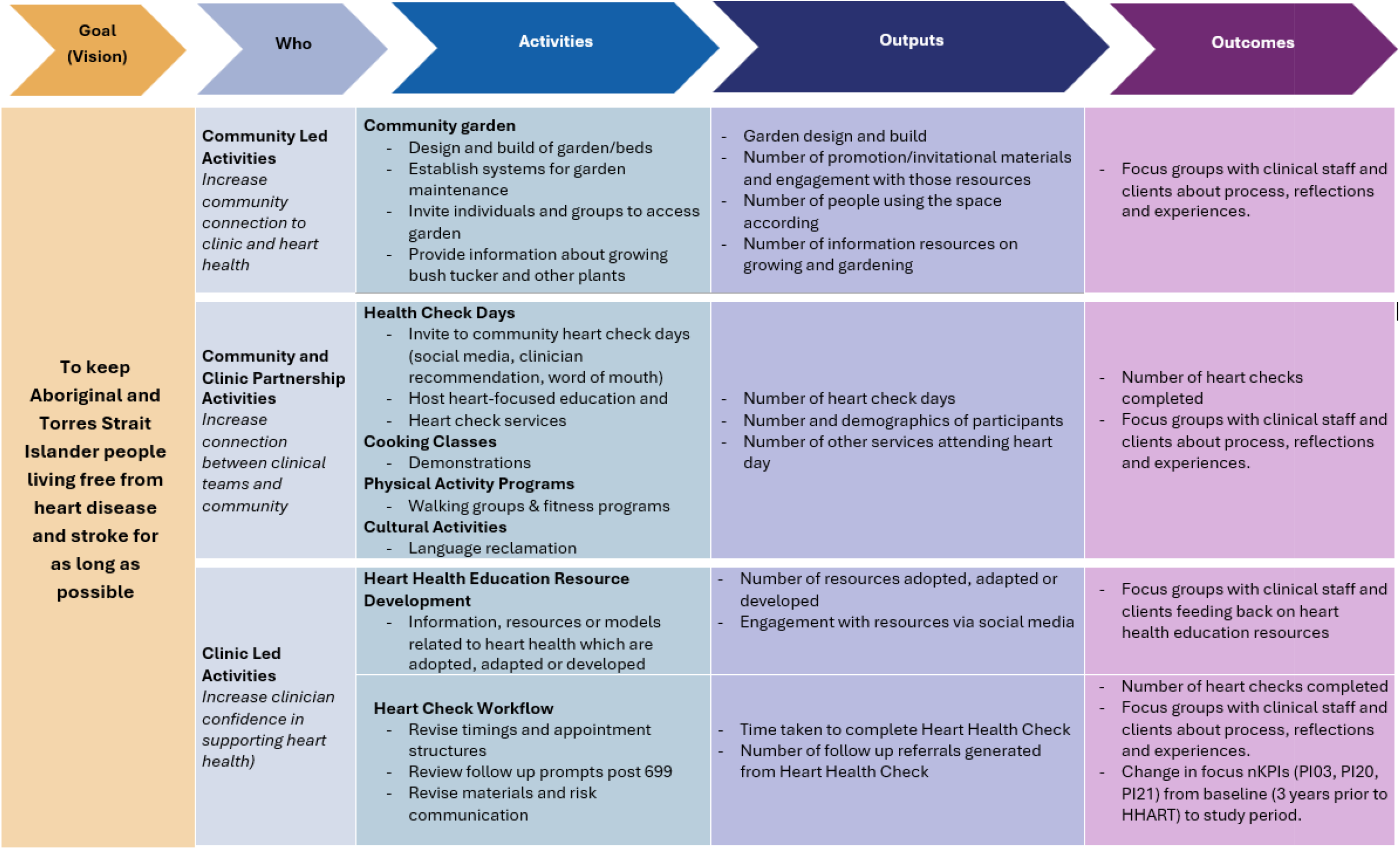
Activity and evaluation plan.

Given the early focus on garden initiatives, we began a systematic review during Phase One titled “Community gardening for the prevention and management of chronic disease with Aboriginal and Torres Strait Islander people in Australia” (PROSPERO 2025 CRD420251138039). Preliminary findings will help inform implementation and evaluation approaches to garden programs in primary care and identify other relevant programs to draw from.

### Strengths and Limitations

A key strength of HHART is its co-designed and culturally respectful partnerships. HHART draws on the expertise of partners to shape strategies that were locally relevant, feasible, and responsive to culture and context. In doing so, Phase One not only generated a clearer understanding of the locally relevant activities and priorities needed to strengthen heart health across community and clinic settings but also established a strong relational and practical foundation for Phase Two implementation. A second key strength of HHART is its multidimensional governance and operational framework, through which Aboriginal and Torres Strait Islander leadership and sovereignty are embedded at all levels of the project. This leadership shapes governance, decision-making, and implementation, strengthening the cultural integrity, accountability, and local relevance of the project while ensuring that co-designed activities are guided by community authority and culturally grounded ways of working. At the service level, each participating site’s activities are supported by the local clinic team, the majority of whom are Aboriginal and Torres Strait Islander staff, together with an Indigenous Community Advisory Group that provides leadership and oversight of co-designed activities within each service. At the project level, Aboriginal and Torres Strait Islander researchers within the investigator team bring professional expertise in heart health and contribute to the design, implementation, and evaluation of the project. Independent academic expertise and strategic research advice are also provided through Thiitu Tharrmay.

One limitation is that the Phase One data was not subjected to formal qualitative analysis. Instead, Phase One was guided by Dadirri approaches, which is collaborative, community-based, and centred on deep listening and the natural flow of conversation. This approach was appropriate to the relational and co-design aims of the project; however, ethics approvals were in place following the initial and rich conversations with partner staff and community members. These conversations were important in shaping shared understandings and informing Phase Two priorities but were synthesised as part of the co-design process rather than analysed through a formal qualitative analytic framework. This is testament to heart health being a priority and a gap with both clinic staff and community members who had mature ideas from the very first conversations, on promoting and protecting the health of Aboriginal and Torres Strait Islander hearts in their communities.

## Conclusion

This study demonstrates the value of co-design as a relational and iterative process for developing culturally meaningful approaches to heart health with Aboriginal and Torres Strait Islander communities. Across the two distinct service contexts, participants consistently framed heart health as embedded in everyday life – shaped by culture, relationships and social conditions – rather than confined to clinical care alone. By bringing together community and clinic perspectives, the findings highlight the importance of strengthening connections between community-led activities and primary health care. The convergence of priorities across sites, particularly community gardening, underscores the potential for locally driven initiatives to support both heart health and broader social and emotional wellbeing. These priorities emerged through processes that centred Aboriginal and Torres Strait Islander knowledges and leadership, demonstrating how community engagement can be operationalised in practice. Phase Two will focus on implementing and evaluating the co-designed activities within each service, including the establishment of community gardens, delivery of Heart Health Days, increasing heart health checks, and the development of culturally relevant education resources. We now seek a third partner organisation to participate in the study, undertaking Phase One. Ongoing partnership with community and clinic stakeholders will guide adaptation, with evaluation examining feasibility, engagement, and sustainability to inform future scale and translation.

## Acknowledgements

We would like to acknowledge and pay our respects to the Traditional Owners of the lands of the Taribelang Bunda, Gooreng Gooreng, Gurang, Bailai and Boandik Peoples, where this research was conducted. We pay our respects to Elders past, present and future. We extend this gratitude to study participants for sharing their views and experiences, the partnering organisations, and HHART Beat for their governance and advice.

## Conflict of interest

The authors declare no conflict of interest.

## Data availability statement

Data from this study are not available due to the sensitive nature of the content discussed.

## Funding

This project was supported by a Medical Research Future Fund Targeted Call for Research on Cardiovascular Health (GNT: 2032711). OP was supported by a National Health and Medical Research Investigator Grant (GNT: 2026852).

## Notes

### Competing Interest Statement

The authors have declared no competing interest.

### Author Declarations

Ethical approval to conduct Phase One was granted by the South Australian Aboriginal Health Research Ethics Committee (AHREC) (#04-24-1160).

## Reference list

1. Mackean, T., et al., Leading the way: the contribution of Aboriginal community controlled health organisations to community health in Australia. Aust J Prim Health, 2025. 31.

2. Dawson, A., et al., Aboriginal community-controlled aged care: principles, practices and actions to integrate with primary health care. Primary Health Care Research & Development, 2021. 22(e50): p. 1–9.

3. Sheppeard, F., et al., The importance of Aboriginal community-controlled organisations, place and the social and emotional wellbeing of frontline workers in south-eastern New South Wales during the COVID-19 pandemic. First Nations Health and Wellbeing - The Lowitja Journal, 2025. 3.

4. Pearson, O., et al., Aboriginal community controlled health organisations address health equity through action on the social determinants of health of Aboriginal and Torres Strait Islander peoples in Australia. BMC public health, 2020. 20(1): p. 1–13.

5. Sinka, V., et al., Chronic disease prevention programs offered by Aboriginal Community Controlled Health Services in New South Wales, Australia. Aust N Z J Public Health, 2021. 45(1): p. 59–64.

6. McGee, M., et al., Mind The Gap, Aboriginal and Torres Strait Islander Cardiovascular Health: A Narrative Review. Heart Lung Circ, 2023. 32(2): p. 136–142.

7. Brown, A., M. Morrissey, and J. Sherwood, Uncovering the determinants of cardiovascular disease among Indigenous people. Ethnicity and Health, 2006. 11(2): p. 191–210.

8. Anand, S.S., et al., Reducing inequalities in cardiovascular disease: focus on marginalized populations considering ethnicity and race. Lancet Reg Health Eur, 2025. 56: p. 101371.

9. Merone, L., et al., Primary Prevention of Cardiovascular Disease in Minority Indigenous Populations: A Systematic Review. Heart Lung Circ, 2020. 29(9): p. 1278–1291.

10. Brown, K., et al., South Australian Aboriginal Heart and Stroke Plan 2022-2027. 2022, Wardliparingga Aboriginal Health Equity, South Australian Health and Medical Research Institute: Adelaide, South Australia.

11. Commonwealth of Australia, National Aboriginal and Torres Strait Islander Health Plan 2021-2031. 2021, Commonwealth of Australia,: Canberra.

12. Commonwealth of Australia, National Strategic Action Plan for Heart Disease and Stroke. 2020, Department of Health: Canberra, Australian Capital Territory.

13. Clare, H., et al., Developing Integrated Healthcare Models for Indigenous People: Insights from a Relational Systematic Scoping Review. J Community Health, 2026. 51(1): p. 124–142.

14. Dawson, A., et al., Ways of working in Aboriginal and Torres Strait Islander Community Controlled Health Organisations: describing a conceptual model of comprehensive primary healthcare characteristics. Aust N Z J Public Health, 2025. 49(4): p. 100267.

15. Campbell, M.A., et al., Contribution of Aboriginal Community-Controlled Health Services to improving Aboriginal health: an evidence review. Aust Health Rev, 2018. 42(2): p. 218–226.

16. Mbuzi, V., P. Fulbrook, and M. Jessup, Effectiveness of programs to promote cardiovascular health of Indigenous Australians: a systematic review. Int J Equity Health, 2018. 17(1): p. 153.

17. Crowe, S., et al., The case study approach. BMC Med Res Methodol, 2011. 11: p. 100.

18. Sibbald, S.L., et al., Continuing to enhance the quality of case study methodology in health services research. Healthc Manage Forum, 2021. 34(5): p. 291–296.

19. Anderson, K., et al., Development of Key Principles and Best Practices for Co-Design in Health with First Nations Australians. Int J Environ Res Public Health, 2022. 20(1).

20. Gerrard, J., et al., Co-design in healthcare with and for First Nations Peoples of the land now known as Australia: a narrative review. Int J Equity Health, 2025. 24(1): p. 2.

21. Indigenous Wellbeing Centre, 2023-24 Annual Impact Report. 2024, Indigenous Wellbeing Centre,: Bundaberg, Queensland.

22. Pangula Mannamurna Aboriginal Corporation. Welcome to Pangula Mannamurna. n.d.; Available from: https://pangula.org.au/.

23. Morey, K., et al., Research ACCORDing to whom? Developing a South Australian Aboriginal and Torres Strait Islander Health Research Accord. First Nations Health and Wellbeing - The Lowitja Journal, 2023. 1.

24. Yardhura Walani. Our Story. 2026 [cited 2026 20 April]; Available from: https://yardhurawalani.com.au/our-story/.

25. Bateman, S., et al., Real Ways of Working Together: co-creating meaningful Aboriginal community consultations to advance kidney care. Aust N Z J Public Health, 2022. 46(5): p. 614–621.

26. Sharmil, H., et al., Participatory Action Research-Dadirri-Ganma, using Yarning: methodology co-design with Aboriginal community members. Int J Equity Health, 2021. 20(1): p. 160.

27. Greenhalgh, T., et al., Achieving research impact through co□creation in community□based health services: literature review and case study. The Milbank Quarterly, 2016. 94(2): p. 392–429.

28. Yadav, U.N., et al., A rapid review to inform the policy and practice for the implementation of chronic disease prevention and management programs for Aboriginal and Torres Strait Islander people in primary care. Health Res Policy Syst, 2024. 22(1): p. 34.

29. Mossop, P., et al., Addressing Social and Cultural Determinants of Health for Aboriginal and Torres Strait Islander People in Chronic Disease Programs: A Scoping Review. Health Promot J Austr, 2025. 36(4): p. e70100.

30. Morey, K., et al., An Aboriginal□led consortium approach to chronic disease action for health equity and holistic wellbeing. Health Promotion Journal of Australia,, 2023. 34(3): p. 634–643.

31. Wali, S., et al., Learning From Our Strengths: Exploring Strategies to Support Heart Health in Indigenous Communities. CJC Open, 2024. 6(7): p. 849–856.

32. Comino, E., et al., A systematic review of interventions to enhance access to best practice primary health care for chronic disease management, prevention and episodic care. BMC Health Serv Res, 2012. 12.

